# Dietary Traits, Systemic Inflammatory Proxies, and Insomnia-Related Outcomes: Exploratory Mendelian Randomization and Population-Based Evidence

**DOI:** 10.64898/2026.07.03.26357235

**Authors:** Yumin Zhou, Yingying Huang, Yu Cao, Xia Bi

**Affiliations:** Shanghai University of Medicine and Health Sciences Affiliated Zhoupu Hospital, Shanghai 201318, China

**Keywords:** insomnia, dietary intake, systemic inflammation, Mendelian randomization, NHANES, CHARLS, population-based analysis

## Abstract

High-dimensional Mendelian randomization (MR) screens can prioritize candidate dietary and immune pathways for insomnia, but their interpretation is constrained by multiple testing, cross-dataset instability, and limited correspondence between genetic constructs and measured population variables. We conducted an exploratory cross-design analysis that combined MR screening of 231 dietary traits and 731 immune phenotypes, targeted cross-release genetic follow-up in FinnGen R12 and R13, and population-based analyses in NHANES and CHARLS. The targeted R13 follow-up prioritised an omelette-related dietary signal (OR 0.773, 95% CI 0.651–0.917; within-layer FDR q=0.00783), a mixed-fruit signal (OR 1.285, 95% CI 1.102–1.498; within-layer FDR q=0.00683), and CD33- and HLA-DR-related immune-cell traits. In NHANES, mapped omelet/scrambled-egg intake was associated with lower odds of sleep problems in 2017-March 2020 (OR 0.746, 95% CI 0.600-0.927; FDR=0.033) and doctor-reported sleep disorder in 2005-2006 (any intake: OR 0.313, 95% CI 0.157-0.624; FDR=0.008; per 50 g: OR 0.721, 95% CI 0.569-0.914; FDR=0.019). Mixed-fruit proxies were not directionally concordant. Higher C-reactive protein (CRP) was associated with sleep problems in NHANES (OR 1.192, 95% CI 1.085-1.309; FDR=0.001) and frequent restless sleep in CHARLS (OR 1.097, 95% CI 1.049-1.147; FDR<0.001). These findings provide exploratory genetic prioritization and population-based association evidence for selected dietary constructs and systemic inflammatory proxies. They do not establish a causal diet-immune-insomnia mechanism, confirm flow-cytometry immune-cell phenotypes, or support dietary intervention recommendations.

## Introduction

Insomnia is a common sleep disorder characterised by persistent difficulty initiating or maintaining sleep together with daytime impairment. It is associated with impaired quality of life, healthcare use, reduced productivity, and cardiometabolic and psychiatric comorbidity [1–4]. Although cognitive-behavioural and pharmacological approaches remain central to management [5], the biological and behavioural heterogeneity of insomnia leaves an important role for identifying modifiable correlates and hypotheses for future testing.

Dietary intake is one candidate domain. Observational studies and reviews have linked sleep duration and sleep quality to dietary pattern, refined carbohydrates, caffeine, alcohol, fatty-acid profiles, and fruit and vegetable consumption [6–9]. Large-scale genome-wide association studies have also identified a polygenic basis for insomnia [10]. However, conventional observational designs are vulnerable to residual confounding, reverse causation, and measurement error. Genetic approaches can complement observational evidence, but they should not be interpreted as a substitute for clinical or experimental confirmation [11–13].

Immune and inflammatory processes offer a biologically plausible bridge between diet and sleep. Sleep and immunity are reciprocally regulated, and immune cells show circadian variation in distribution, activation, and cytokine signalling [14–17]. Specific flow-cytometry phenotypes, including CD33- and HLA-DR-related traits, can provide genetically informative candidates, but they are not directly measured in most population surveys and must therefore be interpreted cautiously [18,19].

Here, we used a cross-design framework to prioritise diet-related and immune-related signals for insomnia-related outcomes. We combined high-dimensional MR screening, targeted cross-release genetic follow-up in FinnGen, and population-based analyses in NHANES and CHARLS. The objective was not to claim clinical replication or a confirmed mechanism, but to evaluate whether selected genetically prioritised signals showed concordant, heterogeneous, or unsupported patterns when mapped to population-level dietary exposures and systemic inflammatory proxies.

## Materials and Methods

### Study design and inference framework

This was an exploratory, cross-design analysis comprising three components: (i) MR screening of dietary traits and immune phenotypes for associations with insomnia liability; (ii) targeted genetic follow-up using FinnGen R12 and R13 insomnia summary statistics; and (iii) population-based analyses of mapped dietary exposures and systemic inflammatory proxies in NHANES and CHARLS (Fig 1). The design intentionally distinguishes genetic prioritisation from population-based association evidence. Neither component directly validates flow-cytometry immune-cell phenotypes, establishes a causal mechanism, or evaluates a dietary intervention.

**Fig 1.**
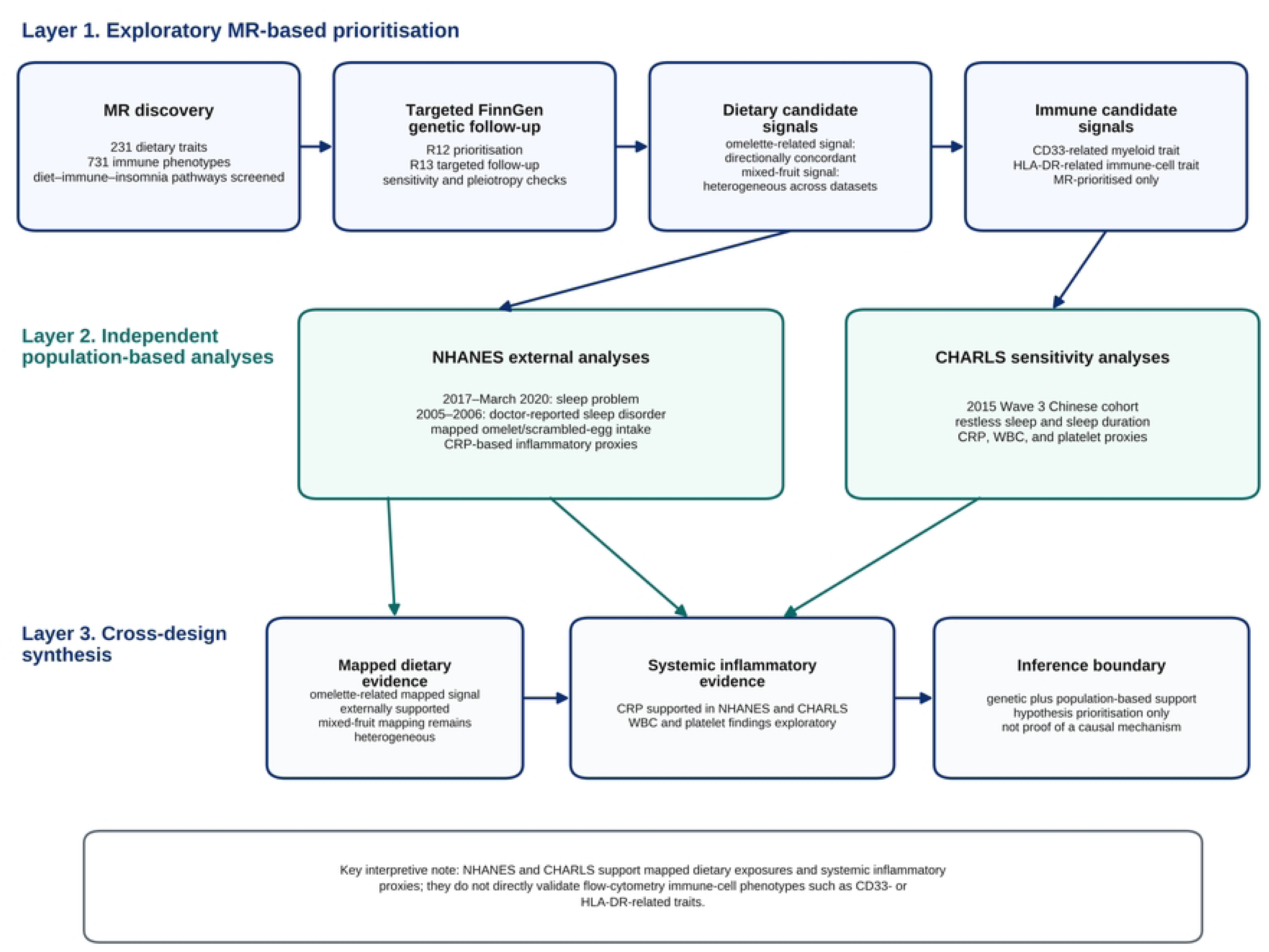
Cross-design study framework. The framework separates exploratory MR screening and targeted cross-release FinnGen follow-up from population-based analyses in NHANES and CHARLS. Population-based analyses of mapped dietary exposures and systemic inflammatory proxies do not directly measure the flow-cytometry immune-cell phenotypes prioritised by MR.

### Genetic data sources and targeted cross-release follow-up

Genetic association data for 231 dietary intake traits were obtained from UK Biobank-derived GWAS resources. Immune-cell phenotype data were obtained from IEU OpenGWAS (accessions ebi-a-GCST90001391 to ebi-a-GCST90002121), including absolute cell counts, median fluorescence intensity traits, surface-antigen levels, morphological parameters, and relative cell-count traits. Discovery insomnia outcome data were derived from Finnish cohorts available through IEU OpenGWAS. FinnGen R12 F5_INSOMNIA was used for cross-release follow-up and included 6,776 cases and 490,763 controls according to the release manifest. FinnGen R13 F5_INSOMNIA summary statistics were then used only for signals that were directionally concordant between discovery and R12. This was targeted genetic follow-up rather than a complete independent re-screening of all dietary and immune phenotypes. FinnGen is described elsewhere [20].

Individual-level data were not available to quantify participant overlap across the discovery, FinnGen R12, and FinnGen R13 sources. The cross-release analyses were therefore interpreted as targeted directional follow-up rather than independent replication, and conclusions were limited accordingly.

### Instrument selection and Mendelian randomization analyses

For dietary and immune phenotypes, SNPs associated with each exposure at P < 1 × 10-5 were selected as candidate instruments to balance instrument availability and statistical power in this high-dimensional exploratory screen. A conventional genome-wide threshold would have substantially reduced available instruments for many behavioural dietary traits. Linkage-disequilibrium clumping used r2 < 0.001 within a 10,000-kb window; palindromic SNPs with ambiguous allele alignment were excluded; and SNPs with F statistics <10 were removed. This pragmatic threshold increases the risk of weak-instrument and pleiotropic bias, so all discovery findings were treated as hypothesis-generating [21].

Analyses used R version 4.2.1 and TwoSampleMR [22]. Inverse-variance weighted (IVW) estimates were the primary results; Wald ratio estimates were used for single-SNP exposures [23]. MR-Egger, weighted median, simple mode, and weighted mode analyses were used where applicable [24–26]. Cochran Q, MR-Egger intercept, MR-PRESSO, and reverse-direction checks were treated as contextual bias-screening analyses [24,27]. Benjamini-Hochberg FDR correction was applied within analytical layers, with FDR-supported findings interpreted separately from nominal exploratory findings [28].

The primary IVW Mendelian randomization estimates were interpreted under the three core instrumental-variable assumptions: (1) relevance, whereby selected genetic instruments were associated with the exposure trait; (2) independence, whereby instrument-outcome associations were not confounded by factors related to insomnia; and (3) exclusion restriction, whereby instruments influenced insomnia only through the exposure. These assumptions cannot be verified directly in this high-dimensional screen. Linkage-disequilibrium clumping, F-statistic filtering, alternative estimators, Cochran Q, MR-Egger intercept, MR-PRESSO, and reverse-direction checks were therefore used as contextual assessments of potential violations.

Because the genetic exposure constructs and the survey-derived dietary and inflammatory measures are not equivalent, the population-based analyses were used only for cross-design association assessment and not as causal validation of the MR estimates.

### Population-based analyses in NHANES

NHANES 2017-March 2020 pre-pandemic data served as the primary population-based analysis, and NHANES 2005-2006 served as an insomnia-focused sensitivity cycle because it included additional sleep-disorder and insomnia-related questionnaire items [29,30]. Adults with required demographic information, sleep outcomes, dietary recall or biomarker data, and non-missing analytic weights were included. The eligible analytic samples comprised up to 7,707 participants in 2017-March 2020 and up to 4,520 participants in 2005-2006, depending on the analysis.

Omelette intake was mapped to omelet/scrambled-egg food-code groups and analysed as any intake and per-50-g intake. Mixed-fruit intake was mapped to available mixed-fruit food codes, with whole-fruit intake used only as a broader contextual proxy. Systemic inflammatory proxies included CRP, white blood cell count, neutrophil count, lymphocyte count, platelet count, neutrophil-to-lymphocyte ratio, systemic immune-inflammation index, and systemic inflammation response index where available. Sleep outcomes included self-reported sleep problems, doctor-reported sleep disorder, diagnosed insomnia in the 2005-2006 cycle, insomnia-related symptoms, and short sleep duration. PHQ-8 was constructed by removing the sleep item from PHQ-9.

Weighted logistic regression was used for binary sleep outcomes and weighted linear models for continuous inflammatory markers. Normalized sample weights were used to avoid treating survey weights as replicated frequencies. Models adjusted for age, sex, race/ethnicity, education, income-to-poverty ratio, body-mass index, smoking, alcohol use, physical activity, diabetes, hypertension, cardiovascular disease, PHQ-8 excluding the sleep item, and total energy intake where appropriate. FDR correction was applied within focused population-based analyses.

For population-based analyses, complete-case models were fitted for the relevant outcome, exposure or inflammatory proxy, covariates, and normalized survey weight; no imputation was performed. Analytic sample sizes therefore varied by outcome and model and are reported in the Supporting Information.

### Population-based analysis in CHARLS

CHARLS 2015 Wave 3 was used to examine whether the inflammatory-proxy layer was observable in a Chinese population [31]. Demographic Background, Health Status and Functioning, Biomarker, Blood,

Weights, and Sample Information files were merged using individual identifiers. Participants aged 45 years or older with available blood-sample weights were eligible; the primary analysis included up to 12,597 participants.

Sleep outcomes included frequent restless sleep based on the CES-D sleep item, short nighttime sleep duration (<6 h and <7 h), and continuous nighttime sleep duration. Inflammatory proxy markers included log(CRP+0.1), white blood cell count, platelet count, CRP >3 mg/L, and the top CRP quartile. Weighted logistic generalized linear models were used for binary outcomes and weighted least-squares models for continuous duration, with normalized blood-sample weights and robust standard errors. Models adjusted for age, sex, rural residence, marital status, body-mass index, smoking, alcohol drinking, hypertension, diabetes, heart disease, stroke, and CES-D score excluding the sleep item.

### Reporting, ethics, and reproducibility

MR reporting was informed by STROBE-MR guidance [32]. This study analysed publicly available summary-level GWAS data and de-identified NHANES and CHARLS data. The relevant publicly available datasets were accessed for research purposes between 18 June 2026 and 30 June 2026. The authors had no access to direct participant identifiers during or after data collection. Ethical approval and informed consent were obtained by the original contributing studies; no new data were collected and no additional institutional review board approval was required for this secondary analysis. The study protocol and analysis plan were not preregistered because the high-dimensional MR screen and cross-design analyses were exploratory. Supporting Information contains study-generated analytic summaries, exposure-mapping metadata, full model outputs, source data for the main population-based figure, and the completed STROBE-MR checklist.

## Results

### Exploratory MR screening and targeted genetic follow-up

The discovery screen identified 11 dietary IVW nominal signals and 28 immune IVW nominal signals; no dietary signal was FDR-supported at the discovery stage. FinnGen R12 identified five dietary and four immune signals that were nominally associated with insomnia and directionally concordant with discovery. These nine candidates proceeded to targeted R13 follow-up.

In FinnGen R13, all 232 requested instrument SNPs for the nine R12-concordant candidates were matched by rsID. Within the dietary candidates, omelette intake retained a protective direction (beta=-0.258, P=0.00313, within-layer FDR q=0.00783; OR=0.773, 95% CI 0.651-0.917) and mixed-fruit intake retained a risk direction (beta=0.251, P=0.00137, within-layer FDR q=0.00683; OR=1.285, 95% CI 1.102-1.498). Within the immune-cell candidates, CD33 on CD66b++ myeloid cells (beta=-0.0139, P=0.00662, within-layer FDR q=0.0244) and HLA-DR on CD33+HLA-DR+CD14- cells (beta=-0.00959, P=0.0122, within-layer FDR q=0.0244) were FDR-supported. These findings provide targeted cross-release genetic prioritisation, not clinical or phenotype-specific biological confirmation (Table 1; Fig 2).

**Fig 2.**
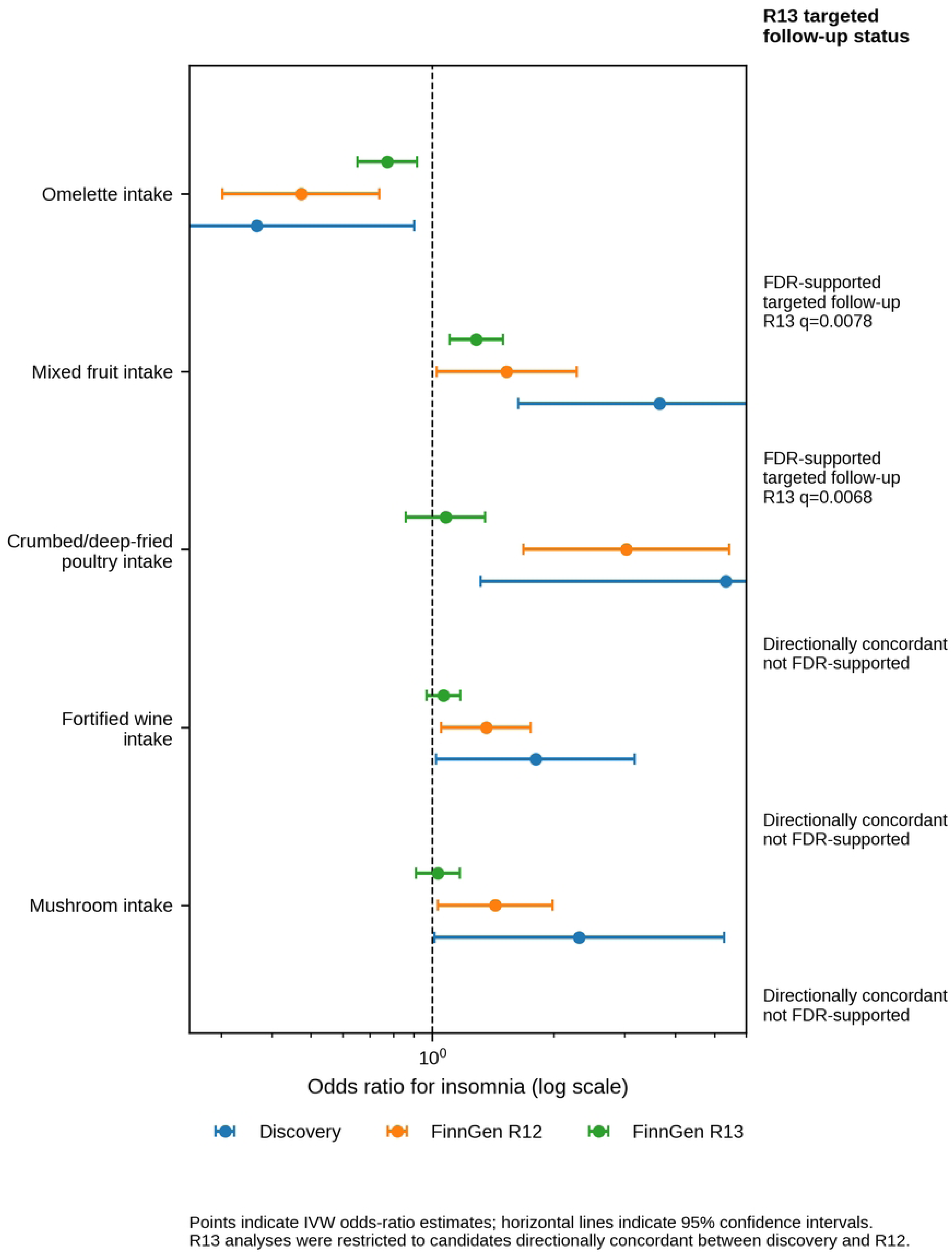
Dietary genetic follow-up. Points indicate inverse-variance weighted odds-ratio estimates and horizontal lines indicate 95% confidence intervals. FinnGen R13 analyses were restricted to dietary candidates directionally concordant between discovery and FinnGen R12; R13 FDR support denotes targeted cross-release genetic follow-up, not clinical or population-based corroboration.

**Table 1.** Analytical framework and inference boundaries.

| Evidence layer | Data and analysis | Primary contribution | What this layer does not establish |
| --- | --- | --- | --- |
| Exploratory MR screen | 231 dietary traits and 731 immune phenotypes; IVW primary estimates; FDR and sensitivity checks. | Prioritises candidate traits for follow-up. | Causality, clinical relevance, or biological confirmation. |
| Targeted FinnGen follow-up | R12 directional prioritisation followed by R13 analyses restricted to R12-concordant candidates. | Assesses cross-release genetic consistency for selected signals. | A complete independent re-screening, patient-cohort evidence, or clinical replication. |
| NHANES population analyses | Mapped dietary exposures, sleep-related outcomes, CRP and blood-count proxies in two survey cycles. | Provides adjusted population-level associations for mapped constructs. | Equivalence of genetic and survey exposure constructs or direct immune-cell measurement. |
| CHARLS population analysis | CRP and blood-count proxies in relation to sleep-related outcomes in a Chinese population. | Assesses whether CRP-related associations are observable in a different population setting. | Direct measurement of CD33/HLA-DR phenotypes or a causal mechanism. |

### Population-based dietary associations

In NHANES 2017-March 2020, any omelet/scrambled-egg intake was associated with lower odds of self-reported sleep problems (OR=0.746, 95% CI 0.600-0.927; FDR=0.033), directionally concordant with the targeted R13 genetic result. In NHANES 2005-2006, any omelet/scrambled-egg intake was associated with lower odds of doctor-reported sleep disorder (OR=0.313, 95% CI 0.157-0.624; FDR=0.008), and the per-50-g analysis was directionally similar (OR=0.721, 95% CI 0.569-0.914; FDR=0.019). Associations with short sleep were not uniformly concordant, so these findings are best interpreted as outcome-specific population associations rather than a replicated dietary intervention effect.

The mixed-fruit candidate was not directionally concordant in NHANES. Whole-fruit and mixed-fruit proxies generally showed protective or inconsistent associations with sleep-related outcomes, whereas the targeted R13 genetic association was in the risk direction. This inconsistency was retained as an example of cross-platform heterogeneity rather than treated as corroborating evidence.

### Systemic inflammatory-proxy associations

In NHANES 2005-2006, higher log(CRP) was associated with higher odds of sleep problems (OR=1.192, 95% CI 1.085-1.309; FDR=0.001). In CHARLS 2015, higher log(CRP+0.1) was associated with greater odds of frequent restless sleep (OR=1.097, 95% CI 1.049-1.147; FDR<0.001). CRP >3 mg/L was also associated with frequent restless sleep (OR=1.167, 95% CI 1.049-1.298; FDR=0.035). White blood cell and platelet findings were weaker and did not support a simple monotonic pro-inflammatory interpretation (Table 2; Fig 3).

**Fig 3.**
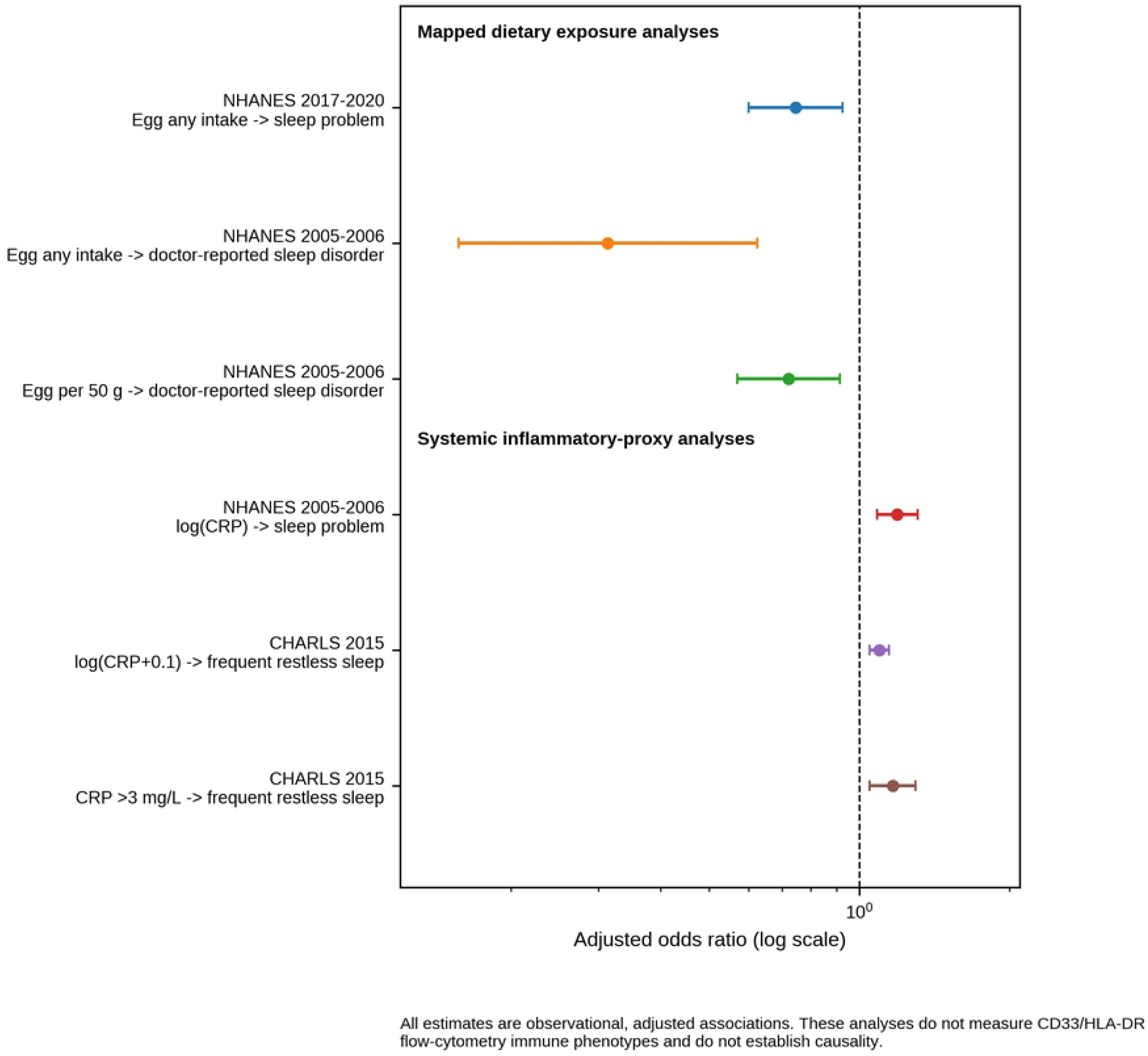
Population-based associations. All estimates are observational, adjusted associations for mapped dietary exposures or systemic inflammatory proxies. These analyses do not measure CD33/HLA-DR flow-cytometry immune phenotypes and do not establish causality.

**Table 2.** Population-based associations selected for the cross-design synthesis.

| Analytic layer | Cohort | Mapped exposure or proxy | Sleep outcome | Adjusted association | Interpretation |
| --- | --- | --- | --- | --- | --- |
| Dietary mapping | NHANES 2017-March 2020 | Omelet/scrambled egg, any intake | Sleep problem | OR=0.746 (95% CI 0.600-0.927); FDR=0.033 | Directionally concordant population association. |
| Dietary mapping | NHANES 2005-2006 | Omelet/scrambled egg, any intake | Doctor-reported sleep disorder | OR=0.313 (95% CI 0.157-0.624); FDR=0.008 | Outcome-specific association in an insomnia-focused survey cycle. |
| Dietary mapping | NHANES 2005-2006 | Omelet/scrambled egg, per 50 g | Doctor-reported sleep disorder | OR=0.721 (95% CI 0.569-0.914); FDR=0.019 | Dose-scaled sensitivity association. |
| Inflammatory proxy | NHANES 2005-2006 | log(CRP), standardized | Sleep problem | OR=1.192 (95% CI 1.085-1.309); FDR=0.001 | Systemic inflammation proxy association. |
| Inflammatory proxy | CHARLS 2015 | log(CRP+0.1), per 1 SD | Frequent restless sleep | OR=1.097 (95% CI 1.049-1.147); FDR<0.001 | Chinese-population association consistent with the NHANES CRP pattern. |
| Inflammatory proxy | CHARLS 2015 | CRP >3 mg/L | Frequent restless sleep | OR=1.167 (95% CI 1.049-1.298); FDR=0.035 | Threshold sensitivity association. |

### Cross-design synthesis and inference boundaries

The population-based analyses supported two limited patterns: an omelet/scrambled-egg association with selected sleep-problem outcomes, and CRP associations with sleep-related outcomes across NHANES and CHARLS. These analyses did not measure the CD33- or HLA-DR-related flow-cytometry immune-cell phenotypes prioritised by MR. They therefore provide population-level association evidence for mapped dietary exposure and systemic inflammatory-proxy layers, not direct phenotypic confirmation of the genetic immune-cell candidates.

## Discussion

This study was designed as a transparent cross-design analysis, not as a mechanistic or interventional study. High-dimensional MR screening and targeted FinnGen follow-up reduced a broad list of candidate traits to a small set of genetically prioritised signals. Population-based analyses then showed partial concordance for a mapped omelet/scrambled-egg exposure and more consistent associations for CRP-related systemic inflammatory proxies. The mixture of concordant and discordant findings is central to the interpretation: it narrows hypotheses but does not turn the results into direct clinical or biological confirmation.

The omelette-related signal was the most consistent dietary candidate. Its direction was protective in targeted R13 follow-up and in selected NHANES sleep-problem and doctor-reported sleep-disorder analyses. Nevertheless, a food-code exposure captured in a single 24-hour recall is not equivalent to a genetically proxied habitual dietary trait. Omelet/scrambled-egg intake may reflect broader meal timing, protein-rich breakfast patterns, socioeconomic context, or overall diet quality rather than an isolated food effect [33,34]. The lack of uniform short-sleep associations further argues against a dietary recommendation based on these results alone.

The mixed-fruit result illustrates the importance of reporting non-concordance. The MR association and targeted R13 follow-up were in a risk direction, whereas NHANES fruit proxies were protective or inconsistent. Differences between mixed-fruit and whole-fruit constructs, sparse exposure mapping, and the contrast between genetically proxied behaviour and short-term recall may all contribute. This candidate should therefore be considered unresolved rather than externally corroborated.

CRP was associated with sleep problems in NHANES and frequent restless sleep in CHARLS, consistent with literature linking sleep disturbance and systemic inflammation [15,17,35]. However, CRP is a broad systemic biomarker and cannot substitute for direct flow-cytometry, single-cell, or experimental measurement of CD33/HLA-DR-related immune-cell phenotypes. The appropriate interpretation is that the population analyses provide a systemic inflammatory-proxy context for the immune layer, rather than evidence for a specific diet-immune-insomnia pathway.

### Limitations

Several limitations are important. First, the MR screen used a relaxed instrument threshold (P < 1 × 10^−5^), which improved coverage of behavioural dietary traits but may increase susceptibility to weak-instrument and pleiotropic bias despite F-statistic filtering and sensitivity checks. Second, FinnGen R13 was a targeted follow-up of R12-concordant candidates rather than a full independent re-screening. Third, the dietary mappings in NHANES were approximate, especially for mixed fruit, and were based on short-term recall rather than habitual intake. Fourth, NHANES and CHARLS outcomes were sleep-related measures rather than standardized clinical insomnia diagnoses in all analyses. Fifth, CRP, blood counts, and derived indices are systemic inflammatory proxies, not direct flow-cytometry measurements of the genetically prioritised immune-cell traits. Finally, all population-based results are observational and cannot establish causality or treatment benefit.

## Conclusions

Exploratory MR screening, targeted FinnGen follow-up, and population-based analyses identified a small set of hypotheses for further study. A mapped omelet/scrambled-egg exposure showed outcome-specific associations with selected NHANES sleep measures, while CRP-related systemic inflammatory proxies were associated with sleep-related outcomes in both NHANES and CHARLS. These findings support future prospective and biological research but do not establish a causal diet-immune-insomnia mechanism, confirm specific immune-cell phenotypes, or justify dietary intervention recommendations.

## Data Availability Statement

All source datasets analysed are publicly available from the original GWAS consortia, IEU OpenGWAS, FinnGen, NHANES, and CHARLS, subject to each resource’s access terms. Study-generated analytical summary tables, variable-mapping metadata, source data underlying Figs 2 and 3 and Table 2, and the completed STROBE-MR checklist are supplied as Supporting Information. Reproducibility materials, including audit utilities, figure-generation scripts, supporting-information materials, and a computational-environment specification, are available through the associated GitHub repository and Zenodo archive. Third-party source datasets are not redistributed; instructions for obtaining the original data resources are provided in the repository documentation.

## Code Availability Statement

Reproducibility materials, including audit utilities, figure-generation scripts, supporting-information materials, and a computational-environment specification, are available at https://github.com/690824069/diet-inflammation-insomnia-reproducibility. An archived release is available at https://doi.org/10.5281/zenodo.20967714. The repository includes instructions for obtaining third-party GWAS, FinnGen, NHANES, and CHARLS resources and for reproducing the deposited study-generated summary tables and figures.

## Supporting Information

S1 File. Supplementary tables and reporting crosswalk. This Excel file contains discovery MR results, cross-release FinnGen R12/R13 follow-up, sensitivity analyses, exploratory mediation outputs, NHANES and CHARLS analysis details, source-data crosswalks, and reporting crosswalk materials.

S3 Checklist. Completed STROBE-MR checklist with manuscript page references and relevant manuscript text.

S2 Data. Source data for Fig 2, Fig 3, and Table 2. This ZIP file contains study-generated numerical source data used to produce the principal figures and the summary population-based association table.

## Author Contributions

Conceptualization: Yumin Zhou, Xia Bi. Methodology: Yumin Zhou, Yingying Huang. Formal analysis: Yumin Zhou. Data curation: Yumin Zhou, Yu Cao. Investigation: Yumin Zhou, Yu Cao. Validation: Yingying Huang, Yu Cao. Visualization: Yumin Zhou. Writing - original draft: Yumin Zhou. Writing - review and editing: Yumin Zhou, Yingying Huang, Yu Cao, Xia Bi. Supervision: Xia Bi. Project administration: Xia Bi. Funding acquisition: Xia Bi. All authors reviewed and approved the final manuscript.

## Ethics Statement

This study analysed publicly available summary-level GWAS data and de-identified NHANES and CHARLS data accessed for research purposes between 18 June 2026 and 30 June 2026. The original studies obtained ethical approval and informed consent. No individual participant was identifiable to the authors, no new data were collected, and no additional ethics approval was required for this secondary analysis.

## Acknowledgments

The authors thank the investigators and participants of UK Biobank, IEU OpenGWAS, FinnGen, NHANES, and CHARLS for making data available for research. We acknowledge the participants and investigators of the FinnGen study.

